# Triage and Referral Behavior Across Patient-Facing Medical AI Products

**DOI:** 10.64898/2026.08.15.26360518

**Authors:** Samuel J. Margolis, Naga Venkata Sai Krishna Maddipatla, Kimon L.H. Ioannides, Lauren E. Wisk, David L. Schriger, Joann G. Elmore

**Author notes:** **Corresponding Author:** Samuel Margolis, BS, David Geffen School of Medicine at UCLA, 885 Tiverton Drive, Los Angeles, CA 90095.

## Abstract

We evaluated nine patient-facing artificial intelligence products using 60 physician-developed standardized clinical cases and 540 multi-turn simulated patient encounters. Although overall triage accuracy showed no statistically significant difference across product categories, referral behavior differed substantially. Branded health AI products more frequently over-triaged low-acuity cases (28% vs 3% vs 2%) and recommended affiliated, fee-requiring clinical services. These findings suggest evaluation of patient-facing medical AI should assess referral behavior alongside overall triage accuracy.

## MAIN TEXT

Patient-facing artificial intelligence (AI) products are increasingly used to evaluate symptoms and inform decisions about where to seek medical care.[1,2] Triage recommendations from these products may influence emergency-department utilization and outpatient clinic visits.[3] Errors in triage recommendations carry potential harm: under-triage may delay necessary treatment, whereas over-triage may expose patients to unnecessary testing, increase health care costs, and worsen clinic and emergency-department crowding.[3]

Evaluation of patient-facing AI products has been limited.[4-8] Large language models’ performance on medical licensing examinations and structured vignettes do not reflect how patients might interact with consumer-facing AI products.[9-11] Prior evaluations relied on single-turn, single-product assessments focused on whether the final answer was correct.[4-7] To our knowledge, few studies have evaluated patient-facing AI products head-to-head using multi-turn standardized-patient encounters, and referral to affiliated clinical services has not been characterized.

We developed a multi-turn patient simulator and evaluated the triage and referral behavior of nine patient-facing AI products across novel clinical cases. We compared branded health AI products, general-purpose consumer chat products, and foundation models to examine how product implementation influences triage behavior.

Across 540 conversations (60 cases × 9 AI products), overall triage accuracy ranged from 81.7% (95% CI, 70–89) to 98.3% (95% CI, 91–100) across the 9 products (**Table 1**). An overall test for a difference in accuracy among the three product categories was not significant (branded health AI products, 88.9% [95% CI, 83–93]; consumer chat products, 90.6% [95% CI, 85–94]; foundation models, 94.4% [95% CI, 90–97]; P = .16).

**Table 1.** Patient-facing AI-Product Healthcare. Triage Performance and Behavioral Indicators

| Patient-facing AI Product | Diagnostic Test Characteristics<sup>□</sup> |  |  |  |  |
| --- | --- | --- | --- | --- | --- |
|  | Over-Triage on Home Cases<br>n/N (%; 95% CI) | Under-Triage on Emergency Cases<br>n/N (%; 95% CI) | Overall Triage Accuracy<br>n/N (%; 95% CI) | Self-Referral<sup>□</sup><br>n/N (%; 95% CI) | Clinical Disclaimer<br>n/N (%; 95% CI) |
| **Branded health AI** |  |  |  |  |  |
| Doctronic | 1/20 (5%, 1–24) | 0/20 (0%, 0–16) | 58/60 (97%, 89–99) | 8/60 (13%, 7–24) | 1/60 (2%, 0–9) |
| PranaDoc | 7/20 (35%, 18–57) | 0/20 (0%, 0–16) | 53/60 (88%, 78–94) | 49/60 (82%, 70–89) | 4/60 (7%, 3–16) |
| Symptomate | 9/20 (45%, 26–66) | 2/20 (10%, 3–30) | 49/60 (82%, 70–89) | N/A | 0/60 (0%, 0–6) |
| *Pooled* | **28% (19–41)** | **3% (1–11)** | **160/180 (89%, 83–93)** | N/A | **5/180 (3%, 1–6)** |
| **Consumer chat products** |  |  |  |  |  |
| ChatGPT | 0/20 (0%, 0–16) | 2/20 (10%, 3–30) | 56/60 (93%, 84–97) | N/A | 9/60 (15%, 8–26) |
| Gemini | 1/20 (5%, 1–24) | 2/20 (10%, 3–30) | 55/60 (92%, 82–96) | N/A | 38/60 (63%, 51–74) |
| Claude AI | 1/20 (5%, 1–24) | 4/20 (20%, 8–42) | 52/60 (87%, 76–93) | N/A | 5/60 (8%, 4–18) |
| *Pooled* | **3% (1–11)** | **13% (7–24)** | **163/180 (91%, 85–94)** | N/A | **52/180 (29%, 23–36)** |
| **Foundation models** |  |  |  |  |  |
| GPT-5.4 | 0/20 (0%, 0–16) | 0/20 (0%, 0–16) | 59/60 (98%, 91–100) | N/A | 1/60 (2%, 0–9) |
| Gemini 3.1 Pro | 1/20 (5%, 1–24) | 0/20 (0%, 0–16) | 58/60 (97%, 89–99) | N/A | 48/60 (80%, 68–88) |
| Claude Opus 4.7 | 0/20 (0%, 0–16) | 1/20 (5%, 1–24) | 53/60 (88%, 78–94) | N/A | 12/60 (20%, 12–32) |
| *Pooled* | **2% (0–9)** | **2% (0–9)** | **170/180 (94%, 90–97)** | N/A | **61/180 (34%, 27–41)** |

Abbreviations: CI, confidence interval; N/A, not applicable.

Despite no statistically significant difference in overall triage accuracy, differences in referral behavior were observed across product categories (**Figure 1; Table 1)**. Branded health AI products over-triaged home-care cases at 28.3% (95% CI, 19–41), compared with 3.3% (95% CI, 1–11) for consumer chat products and 1.7% (95% CI, 0–9) for foundation models (difference vs the other categories, 25.8 percentage points [95% CI, 15–38]; Benjamini-Hochberg–adjusted P < .001; **Table 1**).

**Figure 1.**
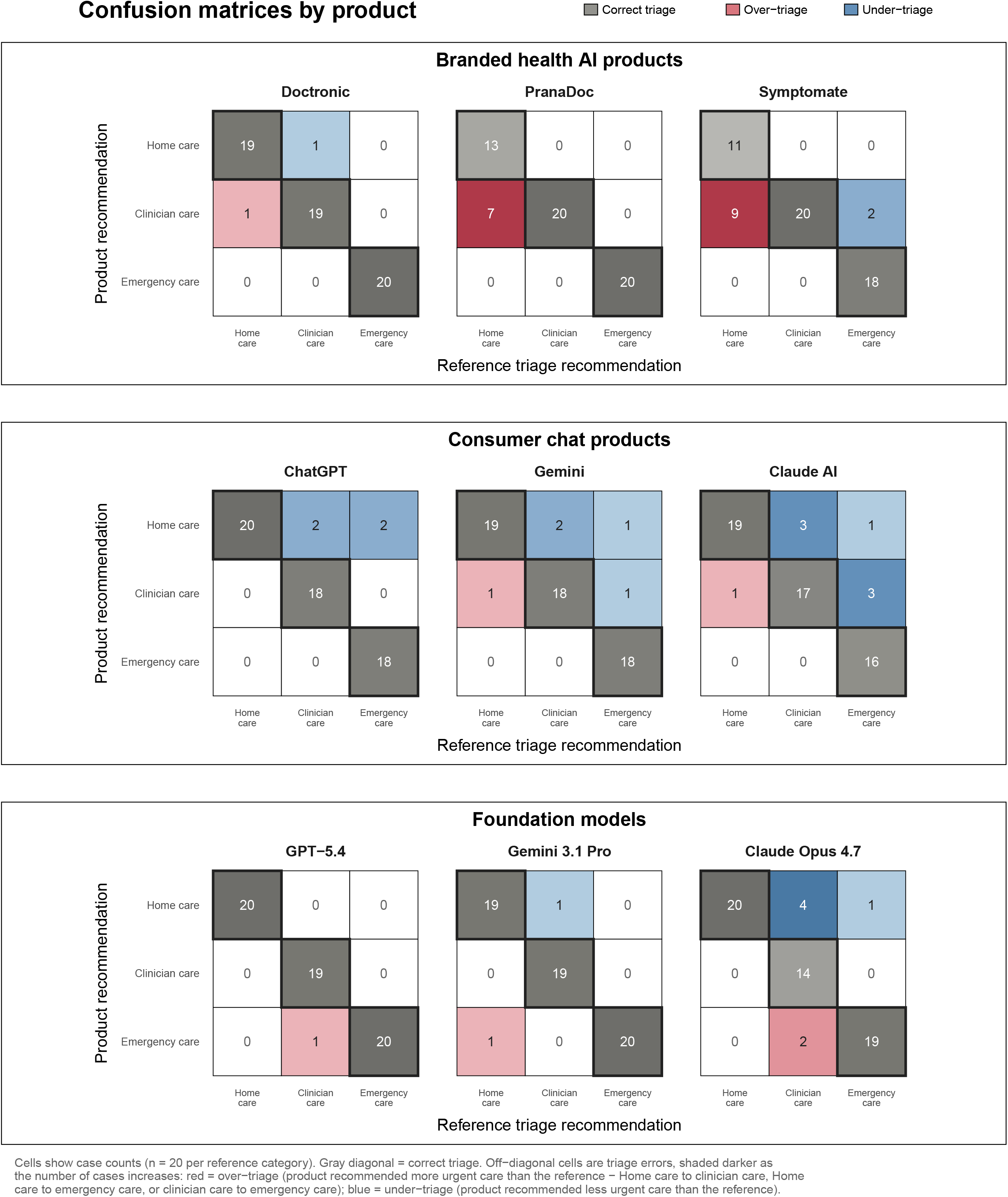
Patient-facing AI Product Triage Accuracy. Product-specific 3 × 3 confusion matrices for each of the nine AI products across 60 standardized cases. Rows indicate the reference (physician-consensus) triage disposition and columns the product’s recommendation, across three tiers: home care, clinician evaluation, and emergency-department evaluation (n = 20 per tier). Cell values show case counts; shading indicates within-row proportions. Diagonal cells are correct triage of the AI product with the reference; cells above the diagonal are over-triage and cells below are under-triage.

Self-referral occurred in the two branded health AI products with affiliated, fee-requiring clinical care: Doctronic 13% (95% CI, 7–24) and PranaDoc 82% (95% CI, 70–89). Self-referral could not occur for Symptomate or products outside the branded health AI category (**Figure 2**).

**Figure 2.**
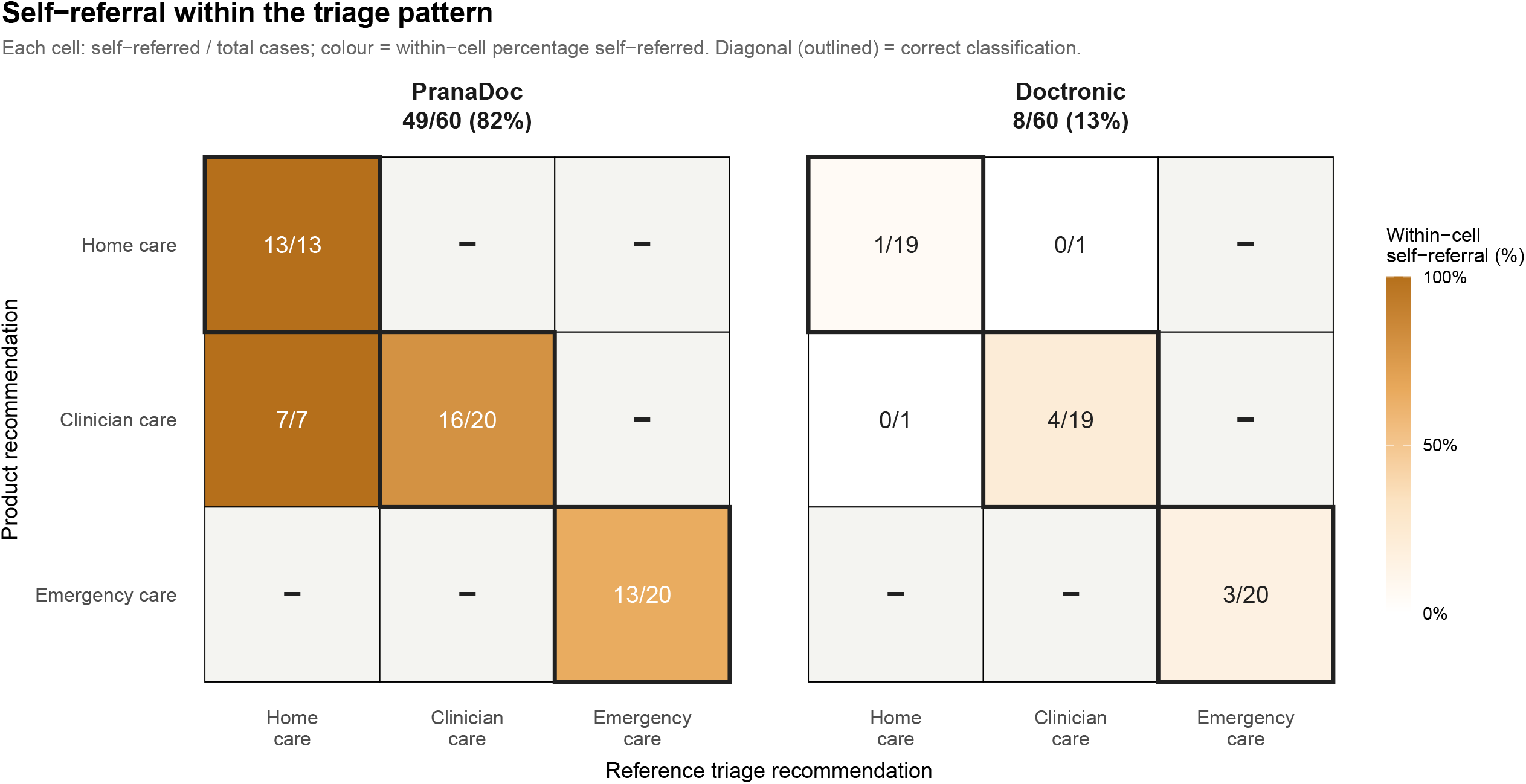
Self-referral of the two branded health AI products with affiliated clinical care. Confusion matrices for the two products offering direct self-referral to affiliated clinical care (PranaDoc, Doctronic). Reference disposition is on the y-axis and product recommendation on the x-axis; each cell reports self-referred cases out of total, shaded by self-referral rate. Symptomate, the three consumer chat products, and the three foundation models are not shown, as they have no affiliated clinical care systems.

Branded health AI products under-triaged 3.3% (95% CI, 1–11) of emergency cases, compared with 13.3% (95% CI, 7–24) for consumer chat products and 1.7% (95% CI, 0–9) for foundation models (consumer chat vs other categories, difference 10.8 percentage points [95% CI, 3–22]; Benjamini-Hochberg–adjusted P = .02). Clinical disclaimers were least frequent in branded health AI conversations (2.8% [95% CI, 1–6]), compared with 28.9% (23–36) for consumer chat products and 33.9% (27–41) for foundation models (difference vs the other categories, −28.6 percentage points [95% CI, −34 to −23]; adjusted P < .001).

Patient-facing AI products have become a front door to medical care. In this evaluation of nine patient-facing AI products, overall triage accuracy showed no statistically significant difference among product categories, but referral behavior differed substantially. Branded health AI products were more likely to direct low-acuity patients toward clinician evaluation and, when available, referred patients to their fee-requiring clinical services. Variation within the branded health AI category was noted, suggesting that referral behavior depends on individual product design. Because these products more often escalated cases that physicians judged appropriate for home care, broader adoption could increase avoidable clinic and emergency-department visits, contributing to crowding and low-value care. Foundation models under-triaged fewer emergency cases than the consumer chat products built on them (1.7% vs 13.3%), and this gap held within each vendor, suggesting that the consumer product’s wrapper, harness, and instructions can reduce the usefulness of triage output relative to the underlying model alone.

Prior evaluations have largely focused on diagnostic, triage, or health-advice accuracy, often using standardized vignettes or single-product assessments.[4-8] We demonstrate that referral behavior is an independent behavioral property that captures clinically meaningful differences between products whose overall accuracy did not differ significantly, and that safety evaluation should assess referral behavior within tiers of triage acuity, including over-triage, under-triage, and self-referral.

This study has limitations. Standardized cases cannot fully capture the ambiguity and longitudinal nature of real clinical encounters. Products were evaluated at a single time point through unauthenticated or free-tier interfaces, and results may not generalize to real-world patient populations, other languages, or paid versions. Presence of a clinical disclaimer was coded from the product’s conversational turns (the body of the encounter), not from any pre-conversation onboarding or click-through screens. We did not evaluate downstream patient behavior or clinical outcomes.

Our findings suggest that patient-facing medical AI should not be evaluated solely on overall diagnostic or triage accuracy. Referral behavior, including systematic over-triage and referral to affiliated clinical services, represents an independent behavioral dimension that may substantially influence healthcare utilization and should become a routine component of evaluation frameworks.

## METHODS

### Study design

This cross-sectional evaluation, conducted May 10, 2026, is reported in accordance with STROBE reporting guidelines.[12] For full details, see the Supplement.

### Cases

Sixty novel clinical case presentations were developed and adjudicated by 3 board-certified physicians across three reference triage tiers (home care, clinician evaluation, and emergency-department evaluation; n = 20 each tier). For each case, the physicians developed a fully specified scenario, including presenting symptoms, relevant history, and predefined standardized answers to questions a product might ask during the encounter. The physicians reached complete agreement on the reference triage recommendation for all 60 cases. Topic selection, structured clinical facts, and full case specifications appear in the Supplement.

### Products evaluated

We evaluated nine AI products across three product categories: (1) branded health AI products (Doctronic, PranaDoc, Symptomate), (2) consumer chat products (ChatGPT, Gemini, and Claude AI), and (3) foundation models (GPT-5.4, Gemini 3.1 Pro, and Claude Opus 4.7). Products were selected to represent the three main routes by which patients currently reach AI-driven triage, choosing widely used options that offered a free or unauthenticated tier within each category.

The consumer chat products and foundation models span the three leading vendors (OpenAI, Google, and Anthropic), allowing each consumer product to be compared with the underlying model from the same vendor. Foundation models were accessed directly through developer interfaces, with no consumer-facing or healthcare-specific layer; consumer chat and branded health AI products add interface, workflow, and safety layers on top of an underlying model. Two branded health AI products are integrated with clinical services that can be recommended to users during the interaction. All products were accessed as unauthenticated or free-tier end users.

### Multi-turn simulated patient encounters

Each case was presented to every product as a multi-turn encounter through a purpose-built patient simulator. The simulator answered only direct questions from the products using predefined case-specific information and never volunteered additional details. Because identical case specifications were used across all products, differences in recommendations reflect product behavior. Each encounter continued until the product stopped seeking information or reached a 30-turn cap, after which a forced-choice prompt required a final triage recommendation.

We release the case set, patient simulator, data-collection harness, coding prompts, and analysis code as TriageBench (see the Data availability and Code availability statements).

### Outcomes

Primary outcomes were (1) over-triage of reference-defined home-care cases to clinician or emergency-department evaluation and (2) self-referral (recommendation of clinical services affiliated with the product on any cases). Exploratory outcomes included under-triage of emergency cases and presence of a clinical disclaimer indicating that the response was not a substitute for medical care. Behavioral indicators (over-triage, under-triage, self-referral, and clinical disclaimer presence) were coded from each case/product conversation using a large language model and validated against physician review in a stratified sample of 30 conversations; agreement was strong across all indicators (Cohen’s κ ≥ 0.97; Supplementary Table S2). Full coding definitions, including how clinical disclaimers were identified, appear in the Supplement.

### Statistical analysis

Proportions are reported with 95% Wilson confidence intervals. [13] For each outcome, proportions for each product category were compared with the pooled proportions of the other two categories using Fisher exact tests with Benjamini-Hochberg correction (two-sided _α_ = .05).[14]

## Data Availability

The standardized case set and de-identified conversation data generated and analyzed in this study will be made publicly available as part of TriageBench in a public repository once final publication in journal is complete.

## Ethics approval

This study did not involve human participant recruitment, intervention, or access to identifiable individual participant data. The University of California, Los Angeles Institutional Review Board determined the study to be exempt (IRB-26-0579). For full details on study methods, see the Supplement.

## Data availability

The 60-case standardized case set and the de-identified conversation data generated and analyzed in this study will be made publicly available as part of TriageBench in a public repository.

## Code availability

The multi-turn patient simulator, data-collection harness, behavioral-coding prompts, and analysis code will be made publicly available as part of TriageBench in a public repository upon acceptance. The benchmark is designed to be extensible: additional products can be evaluated by adding a target adapter, and the rubric and case set can be modified for follow-on hypotheses.

## Author contributions

S.J.M., D.L.S., and J.G.E. conceived and designed the study. S.J.M., K.L.H.I., and D.L.S. developed and adjudicated the clinical cases. S.J.M. and N.V.S.K.M. built the multi-turn patient simulator and data-collection software and acquired the data. S.J.M., N.V.S.K.M., and L.E.W. analyzed and interpreted the data. S.J.M. drafted the manuscript. K.L.H.I., L.E.W., D.L.S., and J.G.E. critically revised the manuscript for important intellectual content. All authors approved the final version.

