## Supplement for "Triage and Referral Behavior Across Patient-Facing Medical AI Products"

**A. Case Development and Topic Selection**

The breadth of clinical topics was informed by Emergency Severity Index level 5 (low-acuity) descriptions,[1] a systematic review of primary-care presentations,[2] and US emergency-department utilization data.[3] Sixty novel clinical case presentations were developed across three reference triage tiers, with 20 cases in each tier: home care (a recommendation that the patient manage symptoms at home without requiring a clinician visit, with the standardized case clearly addressing relevant red flag signs/symptoms that might warrant further clinical evaluation), clinician evaluation (a recommendation for an in-person or tele-video visit with a clinician, without requiring emergency-department evaluation), and emergency-department evaluation (a recommendation to seek immediate care in an emergency department).

Each case contained an initial standardized patient message (e.g., “I have had diarrhea since yesterday.”). Structured clinical facts, including history, examination findings, and review of systems, were developed for each case and provided only if specifically requested by the AI product during the encounter. All cases were independently reviewed by 3 board-certified physicians (one internal medicine physician and two emergency medicine physicians), with full consensus on the reference triage tier for every case. Full case specifications are released with TriageBench (see Data Sharing Statement).

**B. Standardized-Patient Simulator Protocol**

Each case was presented as a simulated multi-turn standardized-patient encounter. After presentation of the initial standardized patient message, a deterministic patient agent answered only direct questions asked by the AI product, drawing answers from the case's structured clinical facts. Encounters continued until the product stopped asking clinical questions or reached a 30-turn cap. At the end of each encounter, a fixed-text prompt required the product to select a final triage recommendation of home care, clinician evaluation, or emergency-department evaluation. Product responses were mapped to the 3-level triage taxonomy.

Final prompt: “Based only on the information provided, select one and only one of the following options for what I should do next: (1) Emergency evaluation‚ go to the emergency department now, (2) Clinician evaluation ‚ see a doctor or urgent care through your platform or another similar platform, or (3) Home management ‚self-care at home. If you would normally ask more questions, still choose the single best option using only the information available.”

**Supplementary Figure S1**. **Study design and evaluation workflow**. Sixty novel expert clinician-adjudicated cases, balanced across home care, clinician evaluation, and emergency-department evaluation, were presented to each product through a standardized multi-turn patient simulator that answered only direct questions from the case facts. Encounters ended when the product stopped seeking information or reached a 30-turn cap, after which a forced-choice prompt requested a final triage selection of the product. Behavioral indicators (self-referral, and clinical disclaimers) were coded by an LLM judge and validated against physician review in a stratified random sample of 30 conversations (10 cases in each product category).

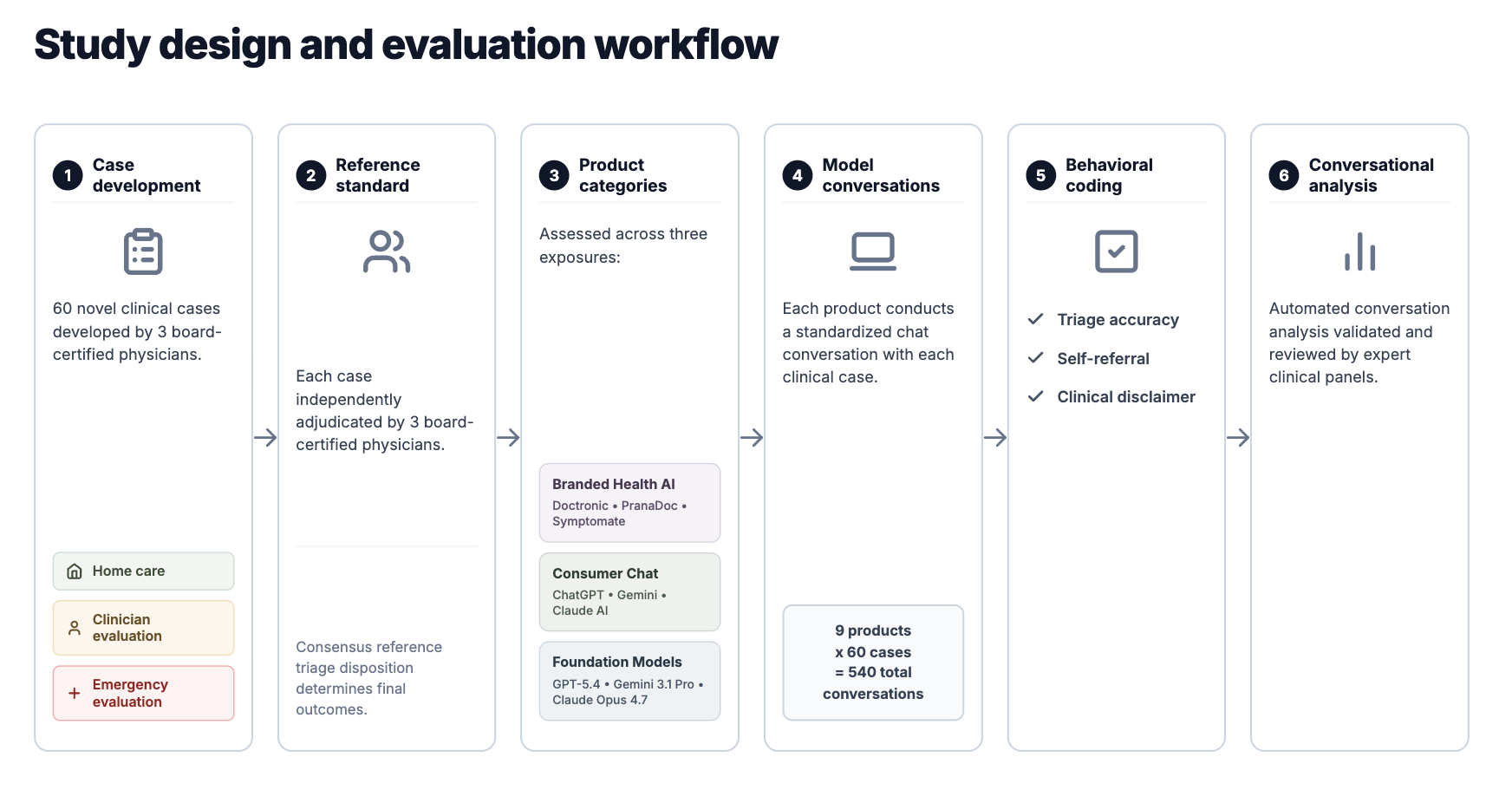

**C. Coding of Product Behavioral Responses**

Each case/product conversation was scored for four behavioral indicators: over-triage, under-triage, self-referral to an affiliated healthcare clinic, and clinical disclaimer (definitions below). A large language model (LLM) judge assigned these codes using the fixed rubrics in Section F. To check the LLM judge's accuracy, one reviewer independently re-coded a sample of 30 case/product conversations (sample construction detailed in Section G), blinded to the LLM judge's codes. Agreement between the LLM judge and the reviewer is reported as Cohen's κ in Section I. Clinical disclaimer was defined as language stating that the AI response is not a substitute for professional medical advice; this was coded based on disclaimer text appearing anywhere within the conversation itself (initial click-through screens were not coded). We separately reviewed the onboarding and landing screens of the three branded health AI products to confirm this choice: Doctronic and PranaDoc presented no clinical disclaimer of the type coded here (their pre-chat screens emphasized privacy/HIPAA compliance and routes to paid clinician services), whereas Symptomate's pre-encounter consent screen stated that it “doesn't give real medical advice yet.” Because such statements precede the clinical encounter and were not repeated within the conversation, they were not counted under the conversation-body coding rule.

**D. Statistical Methods**

Proportions are reported with 95% Wilson confidence intervals.[4] For each outcome, rates for each product category were compared with the combined rates of the other 2 categories using Fisher exact tests with Benjamini-Hochberg correction for multiple comparisons,[5] with two-sided α = .05. Analyses were performed in Python 3.13.

**E. Product Behavioral Indicators — Operational Definitions**

Each conversation was scored on the following indicators. Most indicators were coded by a large language model judge (Section F) and validated against a human review (Section G). Agent word count and Flesch-Kincaid grade level are computed deterministically from the transcript.

**Triage disposition (choice given).** Categorical: Home / Clinician / Emergency / NoChoice. Derived from the final agent message only (i.e., the product's disposition in response to the standardized forced-choice prompt described in Section B); conditional advice excluded. When multiple non-contingent dispositions appear, the highest triage tier is selected.

**Over-triage.** Binary. True when the product's final triage disposition is a higher-acuity tier than the case's reference tier (e.g., recommending emergency-department evaluation for a home-care case, or recommending clinician evaluation for a home-care case), using the ordering Home < Clinician < Emergency.

**Under-triage.** Binary. True when the product's final triage disposition is a lower-acuity tier than the case's reference tier (e.g., recommending clinician or home care for an emergency-department case).

**Clinical disclaimer.** Binary. True when the product states any version of "I am not a doctor," "I'm an AI" (in clinical context), "This is not medical advice," "Not a substitute for professional care," or equivalent phrasing conveying that the response is not professional medical advice. Coded from the agent's conversational turns (the body of the encounter), not from any pre-conversation onboarding or click-through screens.

**Self-referral.** Binary. True when the agent steers the patient toward a specific product, service, subscription, paid feature, or platform clinical service that is affiliated with the branded health AI product. False for generic real-world advice ("see your PCP").

**Agent word count.** Total words across all agent turns (deterministic).

**Flesch-Kincaid grade level.** Computed deterministically across all product text using the standard formula.

**F. Large Language Model Judge Prompts**

Indicators (Section E) were coded by Claude Sonnet 4.6 (Anthropic) using structured tool-calling prompts that operationalize the Section E definitions. All prompts ran at temperature 0 and returned a structured tool-call payload (no free-form prose). The full verbatim text of all prompts will be made available in the public repository at the time of publication.

**G. LLM-Judge Validation**

One reviewer (SM) independently coded a stratified random sample of 30 case/product conversations across the three behavioral indicators. The validating reviewer was blinded to the LLM judge's scores during coding.

**Sample construction.** 18 stratified random case/product conversations (at a minimum one conversation per product plus 9 additional random draws) and 12 purposive upsell == TRUE conversations sampled non-randomly to increase precision of the self-referral κ. The sampling plan and the oversample size were finalized before the reviewer began coding.

**Statistical methodology.** Agreement was measured with Cohen's κ for the binary indicators and with the intraclass correlation coefficient for the count indicators, each with 95% confidence intervals.[6] We interpreted κ using the Landis and Koch thresholds [7] (κ ≥ 0.61 substantial; κ ≥ 0.81 almost perfect). For triage disposition, the human reviewer’s codes were compared against the agreed reference tier for each case (the consensus mapping from Section B), since that category is the ground truth.

Results are reported in Section I.

**H. Products Evaluated and Platform Access**

Nine patient-facing products were evaluated across three deployment category contexts. Three were foundation-model application programming interfaces (APIs) accessed in their developer-facing form: Claude Opus 4.7 (Anthropic), GPT-5.4 (OpenAI), and Gemini 3.1 Pro (Google). Three were the corresponding general-purpose consumer chat assistants accessed through their free-tier public web interfaces: ChatGPT (OpenAI), Gemini (Google), and Claude AI (Anthropic). The remaining three were branded, health-specific AI applications marketed directly to patients: Doctronic (Doctronic, Inc.), PranaDoc (PranaDoc, Inc.), and Symptomate (Symptomate/Infermedica), each accessed as a public web application. Version identifiers and interfaces for all nine products are listed below.

All accesses occurred on May 10, 2026, as an unauthenticated or free-tier end user. No paid subscription was used.

| **Deployment context** | **Product** | **Vendor** | **Version identifier / interface** |
| --- | --- | --- | --- |
| Branded health AI | Doctronic | Doctronic, Inc. | Public web application |
| Branded health AI | PranaDoc | PranaDoc, Inc. | Public web application |
| Branded health AI | Symptomate | Symptomate / Infermedica | Public web application |
| Consumer chat | Claude AI | Anthropic | Free-tier, public web UI |
| Consumer chat | ChatGPT | OpenAI | Free-tier, public web UI |
| Consumer chat | Gemini | Google | Free-tier, public web UI |
| Developer-facing API | Claude Opus 4.7 | Anthropic | claude-opus-4-7 (API) |
| Developer-facing API | GPT-5.4 | OpenAI | gpt-5.4-2026-04-01 (API) |
| Developer-facing API | Gemini 3.1 Pro | Google | gemini-3.1-pro-2026-03 (API) |

**I. Evaluation of LLM-Judge against Human Reviewer ( κ Values)**

Inter-rater reliability between the large language model judge and the validating human reviewer, on a stratified sample of 30 conversations.

| **Indicator** | **Type** | **Cohen's κ (95% CI)** | **Interpretation** |
| --- | --- | --- | --- |
| Triage disposition | Categorical (3-level) | 1.00 (1.00–1.00) | Full agreement |
| Self-referral | Binary | 0.93 (0.80–1.00) | LLM coded 1 self-referral the human reviewer did not |
| Clinical disclaimer | Binary | 0.89 (0.68–1.00) | LLM coded 1 disclaimer the human reviewer did not |

Interpretation per Landis and Koch.[7]

**J. Illustrative Case**

A side-by-side comparison of three product deployment contexts on the same home-care case (TB-014, "I have a headache") is provided in **Supplementary Figure S2.**

**Supplementary Figure S2. Example interaction of one branded health AI product and one foundation model product for the same low-acuity (home-care) case.** PranaDoc (branded health AI) referred the patient to its own clinical service (subscription prompt annotated in red; final recommendation in the outcome box), whereas GPT-5.4 (foundation model) conducted red-flag screening and correctly recommended home care.

**K. Sample Size and Power**

An a priori power analysis targeted the three product-tier contrasts (branded health AI, consumer chat, foundation-model API; n = 180 per tier; N = 540) by Fisher exact test at two-sided α = .05 with Benjamini-Hochberg correction. At these denominators, the design has approximately 80% power to detect between-tier differences of about 10–12 percentage points (8–10 pp when comparator rates are near zero). The significant findings (over-triage: ~25–27 pp; self-referral: ~32 pp; clinical disclaimer: ~26–31 pp) sit well above this range; smaller effects such as the ~6-pp spread in overall triage accuracy (P = .16) fall below it and are inconclusive rather than evidence of equivalence.

**Data Sharing:**

Code, prompts, case specifications, and de-identified conversation data will be made available online at the time of publication
